# Trends and landscape of cardiovascular disease studies characteristics on ClinicalTrials.gov from 2012 to 2021

**DOI:** 10.1101/2023.06.12.23291308

**Authors:** Bharat Rawlley, Kannu Bansal, Utkarsh Dayal, Dhawani Julka, Ishita Salooja, Andres Cordova Sanchez, Kartik Gupta, Sandeep Kumar, Debanik Chaudhuri

## Abstract

**Introduction:** To analyze cardiovascular disease (CVD) studies from the United States registered on ClinicalTrials.gov focusing on characteristics associated with their external validity; the presence of Data Monitoring Committee/Data Safety Monitoring Board (DMC/DSMB), randomization, blinding, and gender of the principal investigators (PIs).

**Methods:** We queried the Application Programming Interface (API) of ClinicalTrials.gov to extract data on characteristics of the identified studies, most notably, DMC/DSMB status, Design Allocation, Design masking, and gender of PIs.

**Results:** We identified 536 studies pertinent to CVD for this analysis. Out of 536, 471 (88%) were interventional studies (Clinical trials) and 65 (12%) were observational studies with interventions. Amongst these, 261 (49%) reported having a DMC, 312 (66%) reported randomization, 224 (48%) reported masking and 122 (19%) of the PIs were women. No trend was seen in the annual proportion of studies with DMC, randomization, masking, and women as PIs (P-trend > 0.05 for all). Multivariable logistic regression analyses were notable for higher odds of DMC (aOR, 5.28; 95% CI, 2.70 – 10.90; P < 0.05) and blinding (aOR, 2.42; 95% CI, 1.29 – 4.64; P < 0.05) in NIH-funded studies and higher odds of being terminated/suspended or withdrawn in basic science studies (aOR, 2.83; 95% CI, 1.07 - 6.90; P < 0.05). No relation was seen between any characteristics and the study being completed.

**Conclusions:** We report on the absence of DMCs, randomization, blinding, women as PIs, and lack of cross-gender collaboration in the leadership of CVD studies without any favorable trend over the past decade. This calls for comprehensive efforts to improve these trends and ultimately, the external validity of studies. We also call for an overhaul of the definition of the phase of a clinical trial that centers around a drug being the intervention.

## Introduction

Clinical studies, especially, clinical trials (CTs) guide clinical practice and are the pinnacle for comparing interventions.^1^ Several studies have demonstrated methodological deficiencies in their registration that lead to the poor external validity of the intervention.^2–5^ While there are multiple factors contributing to the external validity of a study, especially its inclusion and exclusion criteria, here, we focus on the status of a Data Monitoring Committee (DMC) / Data Safety Monitoring Board (DSMB), presence of randomization, masking of investigators and/or participants and gender of the Principal Investigator (PI) as factors associated with external validity of studies. Additionally, we report on how the source of funding, the purpose of the study, the study phase, and the interventions involved relate to the status of DMC, randomization, and masking. Lastly, we investigate if the PIs gender in addition to the erstwhile mentioned characteristics are related to it being completed and being terminated/suspended or withdrawn. We added PIs gender to this additional analysis to investigate if a particular gender faces additional barriers in completing studies. To the best of our knowledge, such a study has not been undertaken in the field of Cardiovascular disease (CVD) in the past decade.

## Methods

This cross-sectional study was a retrospective analysis of a publicly available database and adhered to the Strengthening the Reporting of Observational Studies in Epidemiology (STROBE) guidelines.

## Data source

We queried ClinicalTrials.gov, a registry of studies maintained by the U.S National Library of Medicine where investigators register study proposals and report results. The Application Programming Interface (API) of ClinicalTrials.gov was queried for interventional studies pertinent to CVD, the search strategy was developed using Medical Subject Heading (MeSH) terms relevant to CVD (eTable1). We only included studies registered between 1^st^ January 2012 to 31^st^ December 2021 and having a PI affiliation with a US institution. All studies were manually reviewed and those not relevant to CVD were excluded. We extracted data on the characteristics of the identified studies (Table 1). Our query resulted in both Observational studies with interventions (E.g., Prospective cohort studies, etc.) and Interventional studies/CTs (Figure 1). The PI’s gender was determined using a validated database from Gender-API (https://gender-api.com), which uses an algorithm based on governmental data and manual additions. Only predictions with an accuracy of ≥ 0.8 for the associated gender were accepted, the remaining were determined by searching for the university or other professional profiles of the PI.

**Figure 1:**
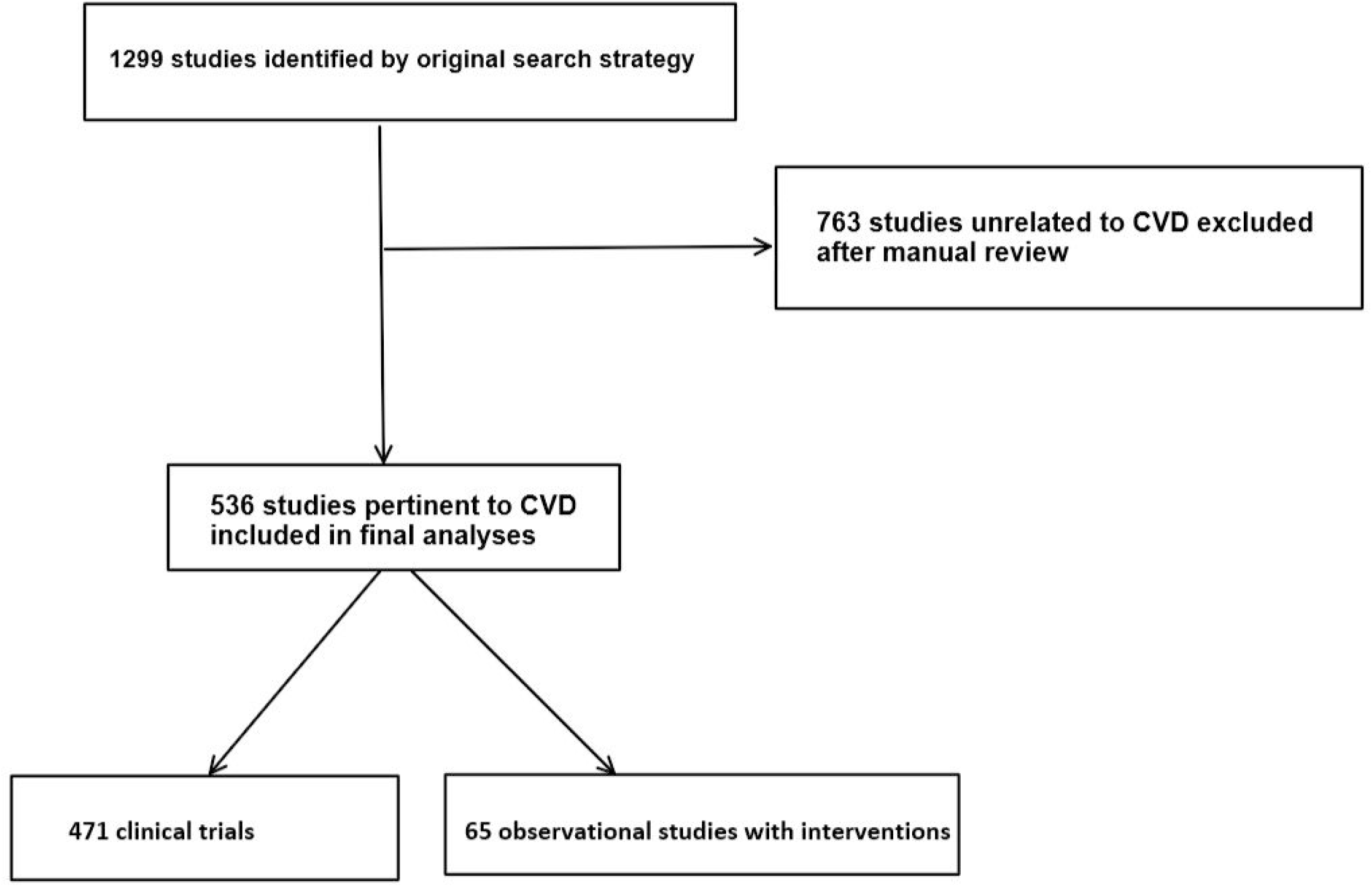
Flow Diagram on selection process of studies included in analyses

**Table 1:**
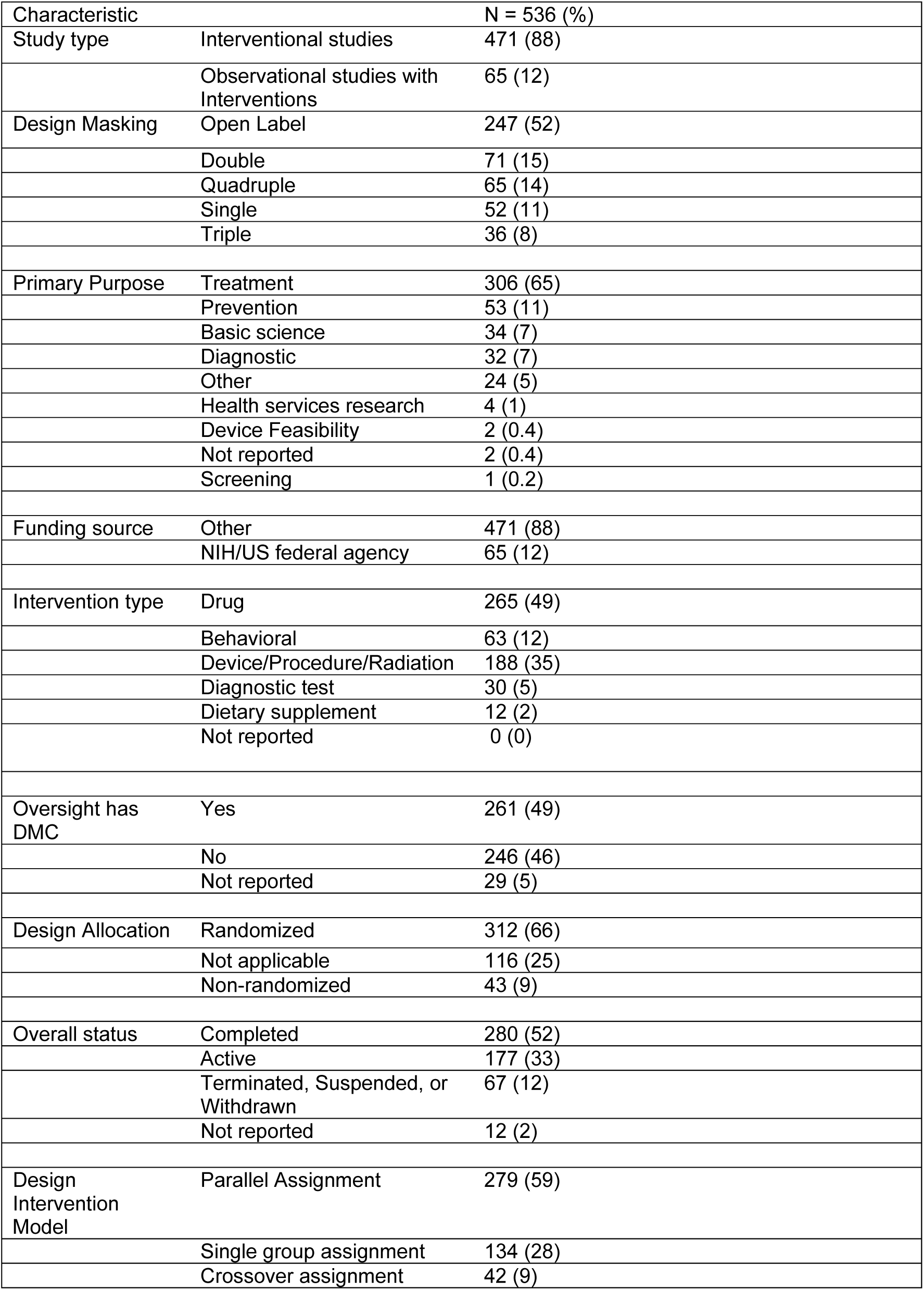
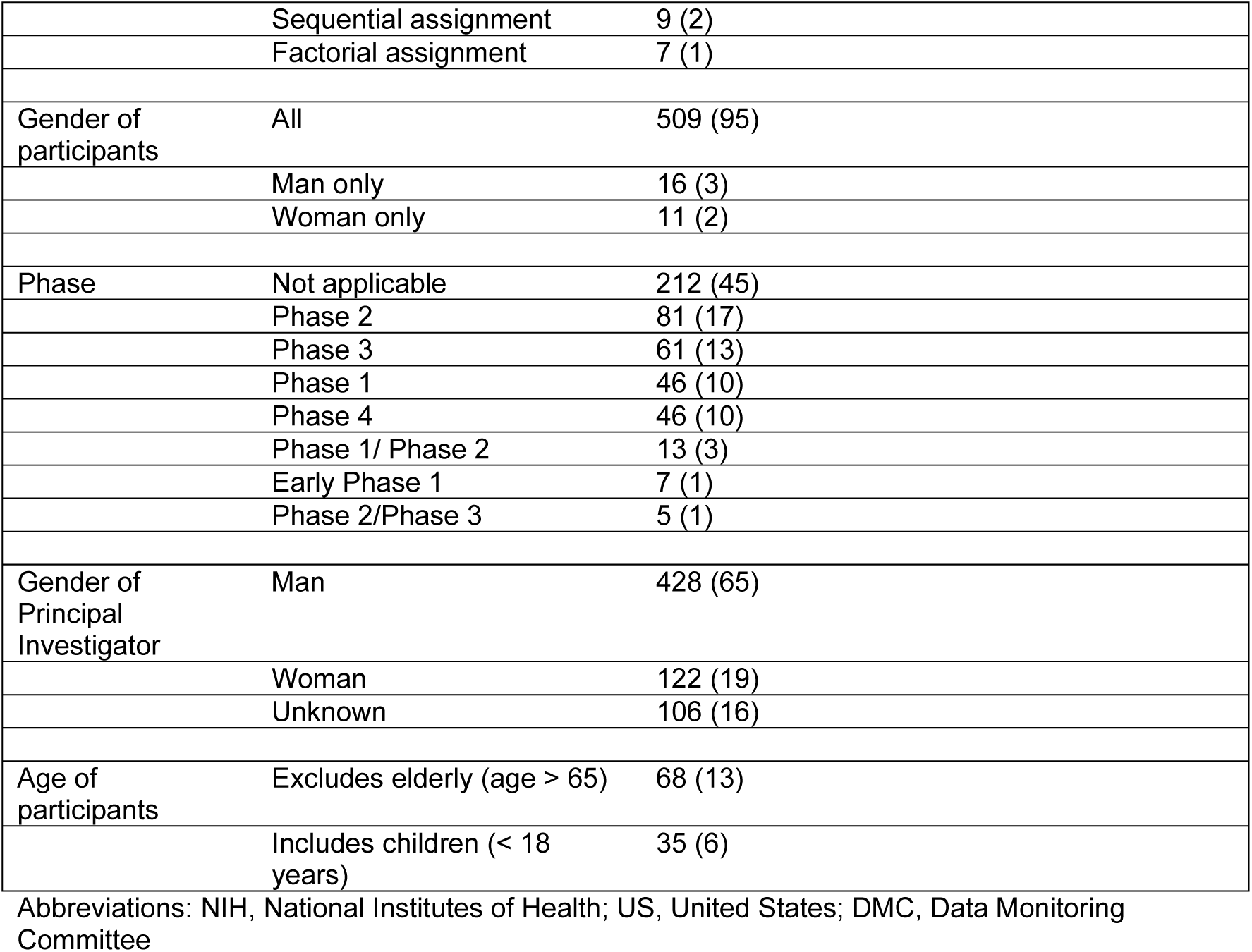
Characteristics of cardiovascular disease studies registered on ClinicalTrials.gov from 1^st^ January 2012 to 31^st^ December 2021.

## Primary outcome measures

Our primary outcome is to illustrate trends in the annual proportion of studies with a DMC, any masking, women as PIs, and randomization as the allocation method from 2012 to 2021. We also report on the type of studies, design masking, primary purpose, funding source, type of intervention, DMC status, allocation design, study status (as of November 2022), design intervention model, gender of the participants, study phase, PIs gender and age of the participants of the included studies (Table 1).

We only included CTs while calculating the proportion of studies belonging to each phase, design intervention model, with masking and reporting randomization as these characteristics do not apply to observational studies with interventions. DMCs apply to observational studies with interventions and hence, they were included in the calculation of the proportion of studies with a DMC. Some studies had multiple PIs and we have taken them all into account while presenting the annual proportion of women as PIs.

## Secondary outcome measures

We use a multivariable logistic regression model to identify study characteristics associated with the presence of DMC, randomization, and blinding. Studies not reporting on the status of DMC were considered as not having a DMC for this analysis. Studies not reporting on the status of randomization or blinding were excluded during their respective regression analyses. Studies reporting that randomization was “not applicable” were also excluded. The dependent variables for these analyses were the purpose of the study, type of intervention, funding source, and study phase. The study phase was categorized into early phase (early Phase 1, Phase 1, Phase 1/Phase 2, Phase 2) or late phase (Phase 2/3, Phase 3 and Phase 4). We selected these dependent variables as findings from this analysis may help regulating bodies formulate guidelines, and interventions directed toward studies of a specific purpose, phase(s), and/or interventions.

Using a different multivariable logistic regression model, we illustrate the relationship between the study characteristics and its overall status; specifically, to recognize characteristics associated with it being completed and being terminated/suspended or withdrawn. The dependent variables for these analyses were the purpose of the study, type of intervention, funding source, study phase, and PI’s gender. We added the PI’s gender to this model as we hypothesized that it may have an impact on the study reaching completion or being terminated/suspended/withdrawn. For studies with multiple PIs, we used the predominant gender group while conducting these analyses. Of note, there were 77 studies with multiple PIs, of which, only 6 studies had a gender heterogenous group of PIs; the rest were led by either all men or all women groups. Two studies were led by an equal number of men and women, and we considered one of them to be led by a man and the other by a woman only for logistic regression analyses.

To report on how study characteristics relate to its reaching completion status, we created a subgroup of studies registered from 2012-2017. We created this subgroup to allow at least 5 years for a study to reach completion status.^6^

## Statistical Analysis

All analysis was conducted using R version 4.1.0 and Microsoft Excel version 2304. The proportion of studies with a DMC, masking, randomization, and women as PIs are presented as percentages stratified by year (Figure 2). A Cox and Stuarts test for trends was performed to report trends in the annual proportion of studies with a DMC, masking, women as PIs, and randomization from 2012 to 2021.

**Figure 2:**
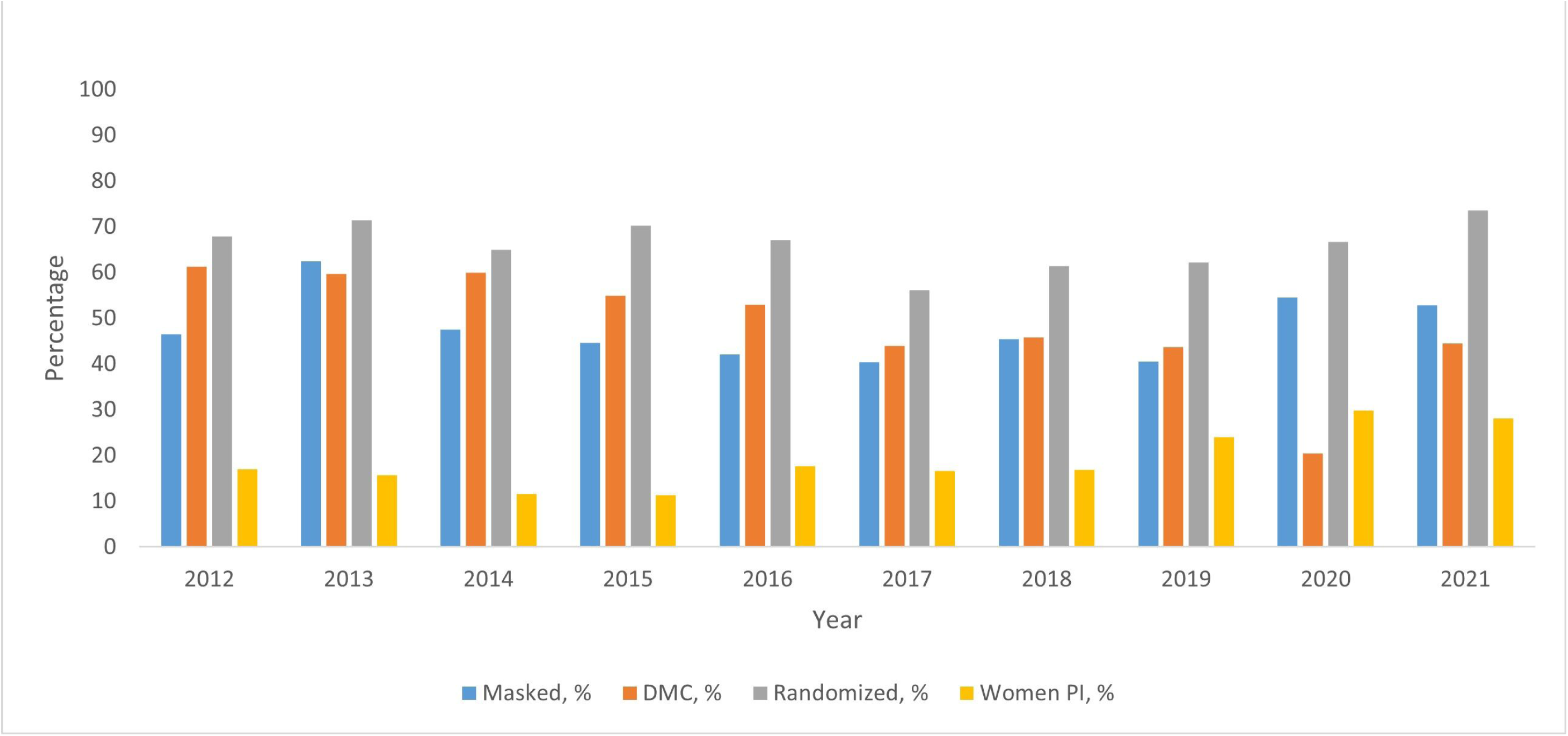
Annual proportion of CVD studies with Masking, DMC, Randomization, and Women as PIs; 2012-2021

A multivariable logistic regression analysis was performed with the presence of DMC, randomization, and blinding as outcome measures. The dependent variables for this analysis are mentioned under secondary outcomes. In a different logistic regression model, we investigate how study characteristics relate to it being completed and being terminated/suspended or withdrawn. The dependent variables for this model have also been described under secondary outcomes.

All statistical tests were two-sided and a P value of < 0.05 was used to designate statistical significance.

## Results

### Trends in the annual proportion of studies with a DMC/DSMB

Among 1299 studies initially identified, 536 were relevant to CVD. Of these, 471 (88%) were CTs and 65 (12%) were observational studies with interventions (Figure 1). Of the 536 studies, 507 (95%) reported their DMC status with 261 (49%) reporting having a DMC. While the annual proportion of studies reporting a DMC decreased from 61% to 44% from 2012 to 2021, the trend was not statistically significant [P-trend = 0.06] (Figure 2).

### Trends in the annual proportion of studies with randomization

All studies reported on their allocation methods, 312 (66%) reported randomization, and 116 (25%) reported that design allocation did not apply to them. The remaining 43 (8%) of the studies were non-randomized. The annual proportion of studies with randomization increased from 68% in 2012 to 73% in 2021, but the trend was not statistically significant [P-trend = 0.37] (Figure 2).

### Trends in the annual proportion of studies with blinding

All studies reported on their masking status, open-label being the most common (247 [52%]) followed by double masking (71 [13%]). The remaining studies reported single, triple, or quadruple masking (Table 1). The annual proportion of studies reporting any kind of masking increased from 46% in 2012 to 53% in 2021, but the trend was not statistically significant [P-trend > 0.99] (Figure 2).

### Trends in the annual proportion of women as PIs and gender distribution of PIs in studies with multiple PIs

There were 656 PIs for the 536 studies in our analysis. We were able to identify the gender of 550 (84%) of the 656 PIs. Of these, 122 (19%) were women (Table 1). There was an increase amongst women as PIs from 17% in 2012 to 28% in 2021, however, this was not statistically significant [P-trend = 0.37] (Figure 2). Additionally, we noted that 77 studies had multiple PIs, of which, only 6 had a gender heterogeneous group, the remaining studies being led by gender homogenous groups; 8 were all women, 61 all men, and in 2 studies we were unable to identify the PI’s gender.

### Characteristics associated with DMC, blinding, randomization, study termination/suspension or withdrawal, and study completion

We performed a logistic regression analysis to identify study characteristics associated with the presence of DMC, Randomization, and blinding. Our results are summarized in Table 2. Some noteworthy observations were that NIH funding was independently associated with the presence of DMCs (aOR, 5.28; 95% CI, 2.70 – 10.90; P < 0.05) and blinding (aOR, 2.42; 95% CI, 1.29 – 4.64; P < 0.05). The odds of all three, that is, DMC, randomization, and blinding were lower in studies with Basic science, diagnostic, and “other” purpose (Table 2).

**Table 2:**
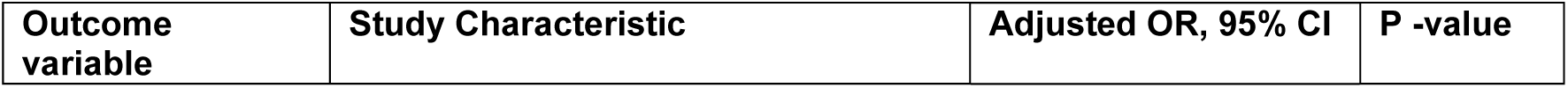
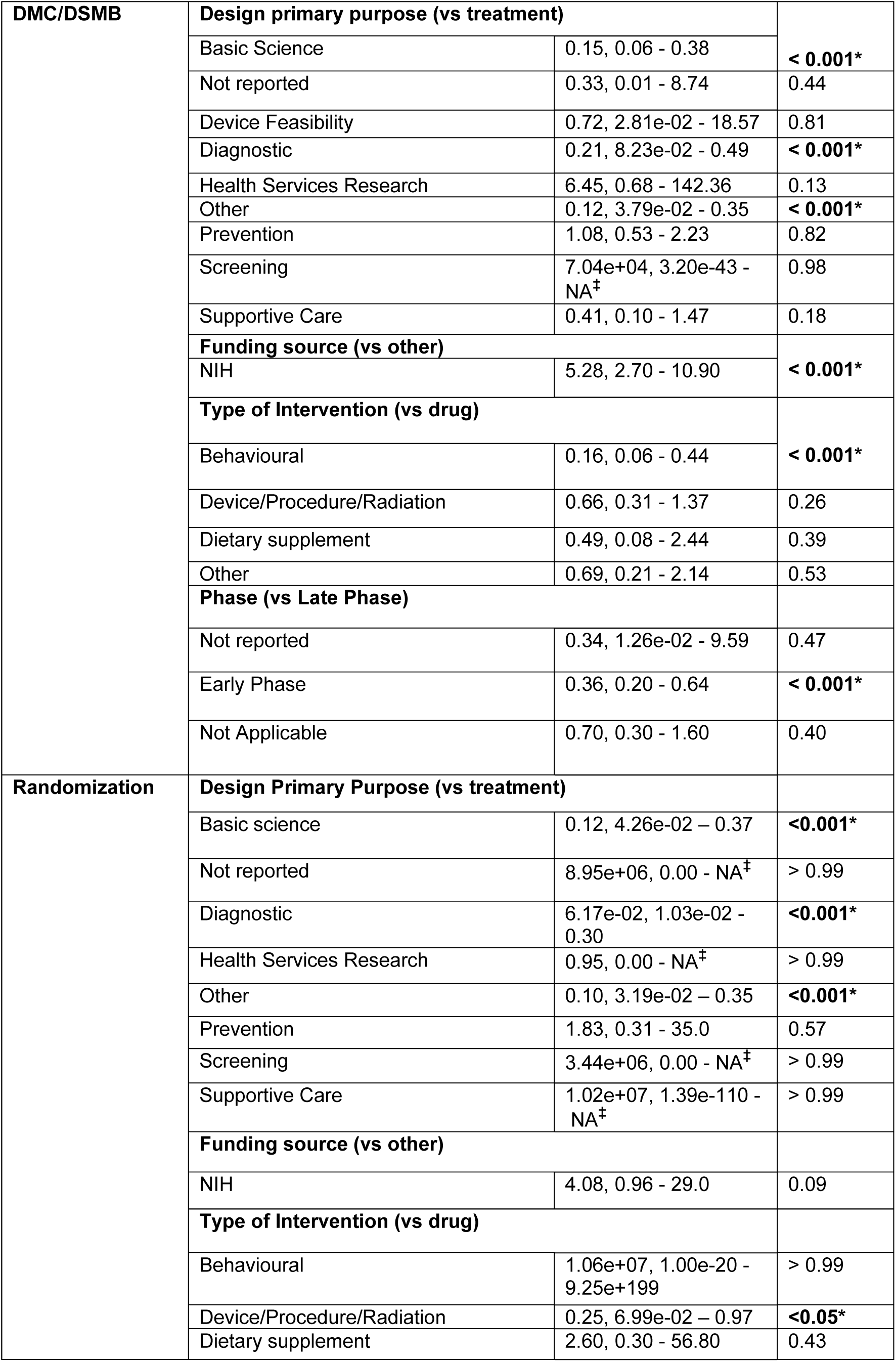
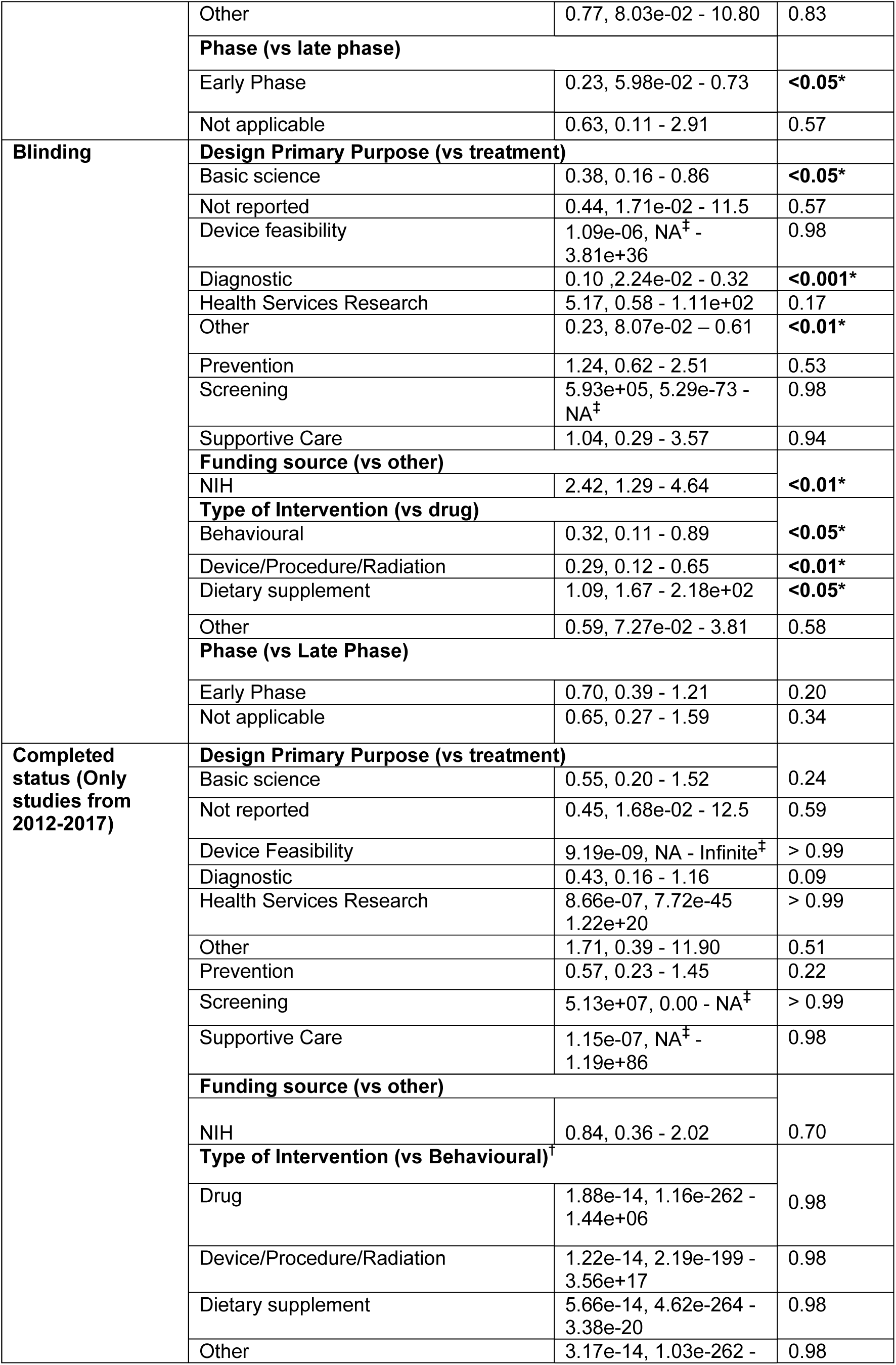
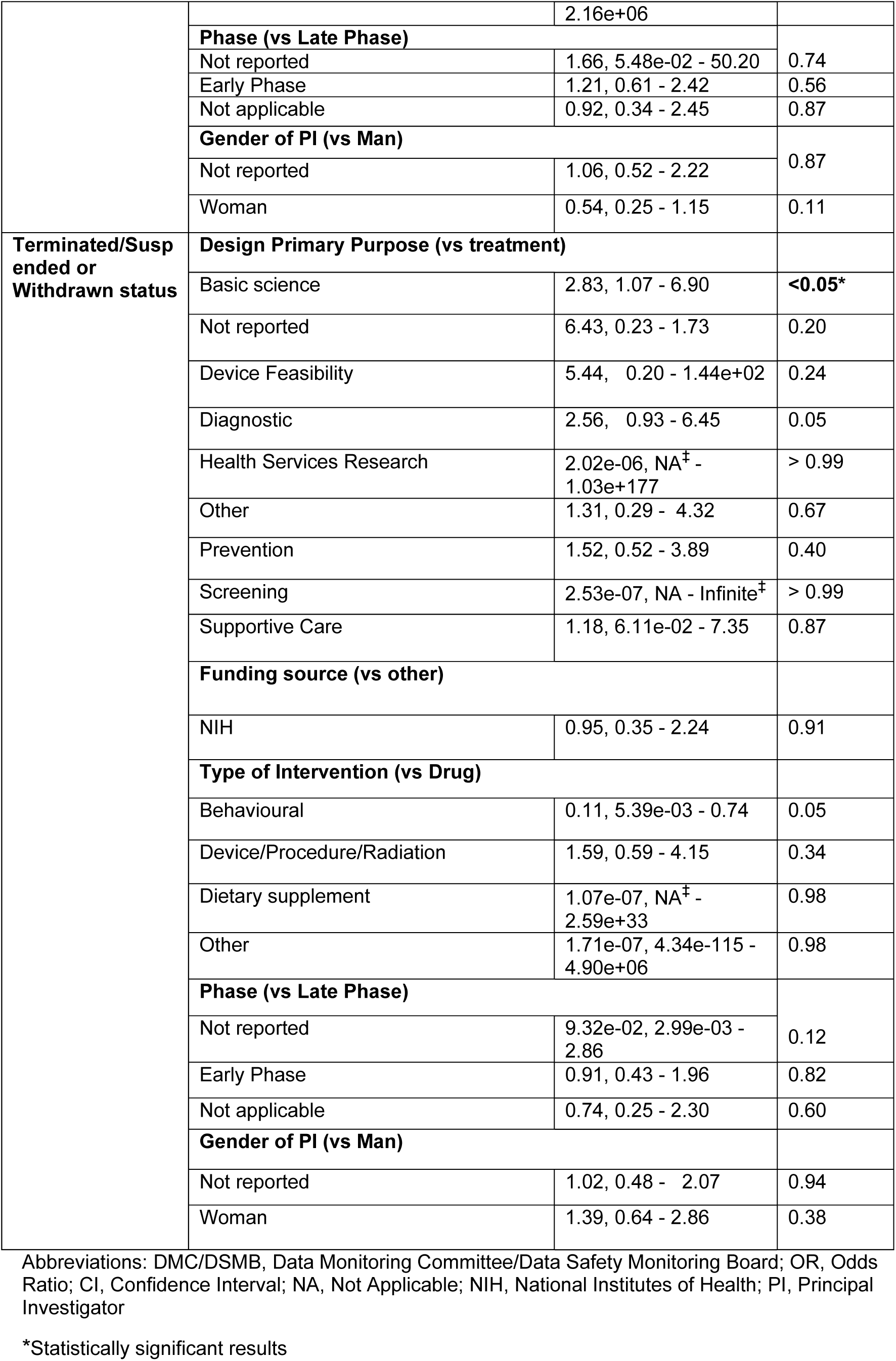
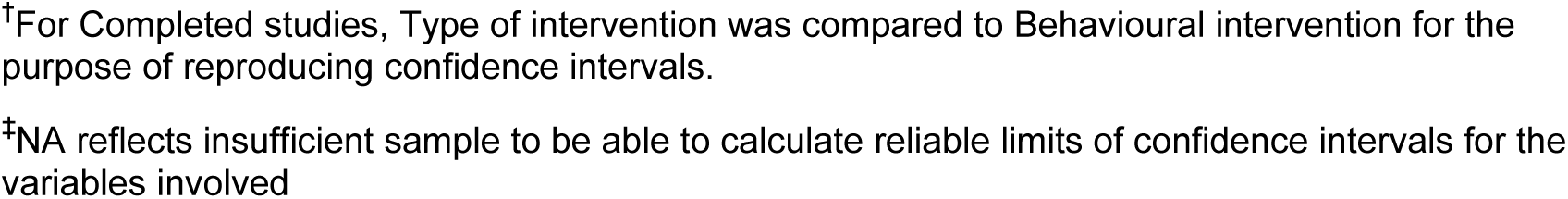
Study characteristics and association with DMC/DSMB, randomization, blinding, study completion status, and termination/suspension or withdrawn status.

In an analysis looking at characteristics associated with studies completion/termination status, the odds of being terminated/suspended or withdrawn were higher in basic science studies (aOR, 2.83; 95% CI, 1.07 - 6.90, P < 0.05) (Table 2). However, none of the study characteristics were associated with it reaching completion status.

## Discussion

In this analyses of 536 studies on CVD, we report that nearly 50% did not have a DMC or any masking. The majority of the studies were randomized (66%) and followed a parallel assignment model (59%). The most common purpose of the study was treatment (65%), and drug was the most common intervention (49%). The study phase was most commonly reported as not applicable (45%) and >50% of the studies reported being completed as of November 2022. Only 19% of the PIs were women and 12% of studies reported being funded by the NIH. No trend was seen in the annual proportion of studies with a DMC, blinding, randomization, or women as PIs and nearly 90% of the studies with multiple PIs were led by gender-homogenous groups. In an analysis relating study characteristics with the presence of DMC, randomization, and blinding, the results were most notable for higher odds of DMC and blinding seen in NIH-funded studies while lower odds of all three, that is, DMC, randomization, and blinding seen in studies with a diagnostic, basic science and “other” purpose. Lastly, on relating study characteristics with their outcome, the odds of being terminated/suspended or withdrawn were higher in basic science studies and no relation was seen between any of the characteristics and the study being completed (Table 2)

A DMC/DSMB is a group of experts who review accumulating data from the study and advise sponsors on its continuation based on evolving understanding of the treatment and scientific merit of the study. While all studies require safety monitoring, not all require a DMC independent from sponsors and investigators. The Food and Drug Administration (FDA) has enlisted multiple criteria where establishing a DMC is recommended. One of them is when invasive interventions are performed or there is a possibility of serious toxicity from the intervention.^7^ We found that 46% of the studies did not have a DMC and 5% did not report on the status of the same. While one may debate the need for a DMC, the lack of a favorable trend calls for improvement, considering that > 80% of the studies in our analyses had a drug/procedure/device or radiation as an intervention.^8^

Randomization reduces selection bias by ensuring that a certain proportion of participants receive each treatment/placebo and the participants being compared have similar baseline characteristics.^9, 10, 11^ We found that 66% of the studies report randomization with no trend over 10 years. In an analysis of CVD CTs from 2007-2010, Califf *et al.* reported 73% had randomization as the allocation method, however, they did not comment on the proportion of trials reporting that a design allocation method did not apply to them.^8^ We have accounted for this in our percentage calculation with 25% reporting that a design allocation method did not apply to them and that may account for the reduction in the proportion of studies reporting randomization in our analyses.

The persistence of gender disparity amongst women as PIs is concerning given that studies led by women have been shown to have greater gender diversity in participants and that increases external validity.^12^ The lack of a trend towards gender parity highlights the deficiencies of the pipeline theory that assumes increasing representation in training would translate into a similar trend as we go to senior roles.^13–15^ Additionally, our finding that only 6 (< 10 %) of the 77 studies with multiple PIs were led by gender heterogeneous groups is concerning given that cross-gender collaboration leads to a diverse thought process which, in turn, drives innovation.^15, 16^ This calls for greater efforts directed towards the inclusion of women as leaders of studies, especially, clinical trials. Some examples are creating flexible funding opportunities and career tracks to continue research productivity during life events like childbirth and adoption that disproportionately impact women.^17, 18^ Lastly, the majority of the studies reported their phase as not applicable, and that reflects studies without an FDA-defined phase. This calls for an overhaul of its definition to include studies with interventions other than drugs. It is noteworthy that 86% of the studies in our analyses with a device/procedure or radiation as intervention had their phase listed as not applicable and that speaks for the need to expand its definition.^19^

Blinding of participants, care providers, and/or investigators is done to reduce bias in the delivery of interventions and assessment of outcome(s).^20^ While blinding indeed reduces systematic bias, it is important to acknowledge the challenges and administrative burden associated with blinding that may paradoxically reduce the generalizability of a study. Analyses assessing subjects’ willingness to enroll have demonstrated an increased willingness to enroll in non-blinded studies. The reduced likelihood of enrollment can lead to a participant population that is less representative of the true patient population, limiting the study’s generalizability.^21^ It is indeed important to assess the advantages and disadvantages of blinding which, in turn, depends on the type of intervention, and the underlying study setting, and then decide if blinding is needed or not. In our analyses, 42% of the studies reported some degree of masking (Table 1) with no trend in the annual proportion of studies reporting masking. Additionally, 46% of the studies were open-label and that may have been intentional for reasons mentioned earlier. Generalizing these findings and calling for an improvement in the proportion of studies reporting any blinding is inappropriate given that this decision is based on the individual study aims and characteristics, however, it is important to report on these metrics given their impact on the validity of the study.

On relating study characteristics to their outcome, we found that studies categorizing their primary purpose as basic science were independently associated with it being terminated/suspended or withdrawn. While COVID-19 may have played a role in this, it has impacted research at all levels and it merits further investigation to understand why studies in the basic science category were disproportionately affected as these are important in understanding the physiology and biomechanics behind an intervention.^22^ We did not find a relationship between any of the characteristics and the study reporting a completion status.

## Limitations

There are several limitations to our analyses. First, ClinicalTrials.gov does not account for all studies performed worldwide as there are other platforms for registering protocols.^23^ Second, studies may have been missed by our search if keywords in our search strategy were not enlisted in the conditions being studied. However, misclassification bias is minimal given that all studies that resulted in the initial search were manually reviewed and excluded if they were unrelated to CVD. Third, while 65% of the studies in our analysis had reached their outcome, 33% were still active/recruiting. Although we only included studies registered from 2012-2017 in the analysis relating study characteristics with their completion, the results from this analysis may change when the remaining 33% reach their result. Lastly, given the limitation of the gender-determining algorithm used by Gender-API, we have reported gender in binary categories of men and women, not considering the full spectrum of the gender identity of an individual.

## Conclusion

Our study reports on the absence of DMCs/DSMBs, randomization, blinding, and women as PIs in CVD studies on ClinicalTrials.gov without any trend over the past decade. Additionally, we report on the lack of cross-gender collaboration in the leadership of studies with multiple PIs. We report on study characteristics associated with the presence of DMC, randomization, blinding, and characteristics associated with study completion or termination. Lastly, we call for an overhaul of the definition of “Phase of Clinical Trial” which, at present centers around a drug being the intervention.

## Data Availability

The data that support the findings of this study are available from the corresponding author upon reasonable request.

## Corresponding Author

Debanik Chaudhuri, MD, Department of Cardiology, 5th floor Upstate Health Care Center, 90 Presidential Plaza, Syracuse, NY 13202

## Author Contributions

Dr. Rawlley and Dr. Chaudhuri had full access to all the data in the study and take responsibility for the integrity of the data and the accuracy of the data analysis.

## Concept and design

Rawlley, Bansal, Dayal, Julka, Salooja, Sanchez, Gupta, Kumar, and Chaudhuri

## Acquisition, analysis, or interpretation of data

Rawlley, Bansal, Dayal, Julka, Salooja, Sanchez, Gupta, Kumar, and Chaudhuri

## Drafting of the manuscript

Rawlley, Bansal, Dayal, Gupta, and Chaudhuri

## Critical revision of the manuscript for important intellectual content

Rawlley, Gupta, and Chaudhuri

## Statistical analysis

Rawlley

## Supervision

Chaudhuri

## Conflict of Interest Disclosures

None

## Funding/Support

None

## Meeting presentation

Parts of this manuscript have been accepted as a poster presentation at the AHA Basic Cardiovascular Sciences 2023 Conference

## Notes

### Competing Interest Statement

The authors have declared no competing interest.

### Clinical Trial

Not Applicable

